# Bacterial strain sharing between humans, animals, and the environment among urban households

**DOI:** 10.1101/2024.08.05.24311509

**Authors:** Daehyun D. Kim, Jenna M. Swarthout, Colin J. Worby, Benard Chieng, John Mboya, Ashlee M. Earl, Sammy M. Njenga, Amy J. Pickering

## Abstract

Identifying bacterial transmission pathways is crucial to inform strategies aimed at curbing the spread of pathogenic and antibiotic-resistant bacteria, especially in rapidly urbanizing low– and middle-income countries. In this study, we assessed bacterial strain-sharing and dissemination of antibiotic resistance across humans, domesticated poultry, canines, household soil, and drinking water in urban informal settlements in Nairobi, Kenya. We collected 321 samples from 50 households and performed Pooling Isolated Colonies-seq (PIC-seq) by sequencing pools of up to five *Escherichia coli* colonies per sample to capture strain diversity, strain-sharing patterns, and overlap of antibiotic-resistant genes (ARGs). Bacterial strains isolated from the household environment carried clinically relevant ARGs, reinforcing the role of the environment in antibiotic resistance dissemination. Strain-sharing rates and resistome similarities across sample types were strongly correlated within households, suggesting clonal spread of bacteria is a main driver of dissemination of ARGs in the domestic urban environment. Within households, *E. coli* strain-sharing was rare between humans and animals but more frequent between humans and drinking water. *E. coli* contamination in stored drinking water was also associated with higher strain-sharing between humans in the same household. Our study demonstrates that contaminated drinking water facilitates human to human strain sharing and water treatment can disrupt transmission.

## Introduction

Bacterial enteric infections cause 9.2% of annual deaths worldwide in children under five.^1,2^ Bacterial infections can trigger acute gastrointestinal illness, while asymptomatic infections are associated with child growth faltering.^3,4^ The incidence and disease burden of bacterial infections are highest in rapidly urbanizing low– and middle-income countries (LMICs),^5^ where over 50% of the urban population lives in informal urban settings with limited access to safe drinking water, sanitation, and appropriate hygiene (WASH) infrastructure.^6^ Furthermore, in these informal urban settings, families often engage in small-scale animal husbandry within confined spaces, increasing the risk of zoonosis.^7,8^ The high prevalence of infectious diseases in urban informal settlements, coupled with a lack of regulated antibiotic use, promotes both the emergence of new antibiotic resistance gene (ARGs) variants through the proliferation of antibiotic resistant bacteria and the spread of antibiotic resistance between bacterial strains via horizontal gene transfer.^6,9^ A recent meta-analysis of fecal metagenomes from 26 countries^10^ identified a higher abundance of ARGs in urban areas in Africa and South-East Asia compared to rural areas.

Enteric pathogens are transmitted to humans by direct contact with infected hosts or via exposure to fecally-contaminated environments.^11^ Exposure to animals has recently garnered attention as a risk factor for enteric infections in LMICs with reports of strain-sharing between humans and domestic or wild animals that are in close proximity.^12,13^ The previous studies have mainly focused on examining bacterial strain sharing between humans and animals,^14,15^ without investigating the environmental exposure pathways through which transmission occurs.^16^ However, humans are regularly exposed to environmental microbiota through consuming drinking water, hand-to-mouth contact with soil, and inhaling soil dust.^7,14^ Understanding transmission pathways across humans, animals, and the environment, along with their spatial range, is important to determine how bacterial transmission could be interrupted and the scale at which interventions need to be implemented.^17^

The epidemiological study of bacterial transmission has traditionally been conducted by sequencing whole genomes of individual isolates selectively cultured from each host.^19^ Sequencing individual isolates has the advantage that it can facilitate the comparison of complete genomes at high-resolution, using metrics such as core genome multilocus sequence typing (cgMLST)^20^ or core genome single nucleotide polymorphisms (cgSNP).^21^ However, due to the labor and cost burden of culturing and sequencing isolates, studies often include only one isolate per sample, potentially missing a diverse population.^7,12^ Sequencing of DNA extracted directly – metagenomics – can describe the diversity of complex microbial communities without a culture step. Several software tools have been recently developed for tracking strains within metagenomes even when they are present at low relative abundance within a complex community.^22,23^ However, reconstructing accurate sequences of ARGs and placing them within their genomic context is often challenging due to low sequencing coverage and high heterogeneity, which can confound assembly.^24,25^ In addition, ARGs are often carried on plasmids, which makes linking these ARGs to their host organisms challenging.^26^ Plate-sweep metagenomics is a recent intermediate approach between isolated colonies and metagenomics that includes a culture step to select for target viable organisms of interest from a sample followed by sequencing the total biomass from the plate. While the overall microbial complexity of a sample is reduced, sequencing effort can still be wasted as most selective media is not species specific.^18,27^

In this study, we aimed to elucidate bacterial strain-sharing and ARG dissemination between humans, domesticated animals, and the household environment. Given its ubiquity and role as a common human commensal as well as clinically important and often antibiotic resistant pathogen, we selected *Escherichia coli* as the target organism.^28,29^ In order to capture strain diversity within each sample and achieve high depth of sequencing coverage, we pooled up to 5 presumptive *E. coli* isolates cultured from the same sample prior to sequencing. Herein, we call our approach “PIC-seq” (Pooling Isolated Colonies-seq). Leveraging matched sample sets of human stool, poultry cloacal swabs, canine feces, stored drinking water, and soil from 50 households in low-income informal settlements in Nairobi, Kenya, we used PIC-seq to determine strain sharing patterns. In addition, we reconstructed the resistomes of pooled *E. coli* isolates at the sample level to explore the role of strain dissemination *versus* horizontal gene transfer in the spread of ARGs.

## Results

### Characteristics of enrolled households

Poultry-owning households (n=50) were enrolled in the study across two subcounties (Dagoretti South: n=25 and Kibera: n=25) in Nairobi, Kenya. In Kibera, all households collected drinking water from chlorinated piped water, as confirmed by chlorine residual testing at the tap during the study. In contrast, water sources in Dagoretti South varied, with households collecting water from boreholes (44%; 11/25 households), piped water (44%; 11/25 households), and water tanks (12%; 3/25 households), none of which underwent chlorination (Supplementary Table 1). Additional household-level water treatment of the collected water was slightly more common in Kibera (24%; 6/25 households) than in Dagoretti South (12%; 3/25 households).

Households from Dagoretti South and Kibera had similar wealth indices, as determined by household assets including availability of electricity, TV, mobile phone, and a stove (Supplementary Table 2). Urban informal settlements in Kenya are often organized into compounds consisting of households sharing a common courtyard, which may include a shared latrine. Households in Dagoretti South were more likely to have latrines inside their compound (76%; 19/25 households) compared to Kibera (20%; 5/25 households), where the majority relied on public facilities located >5 meters away outside of the household compound (64%; 16/24 households) (Supplementary Table 3). The purpose for poultry ownership across both subcounties was primarily nutrient provision via meat (Dagoretti South: 88%, 22/25 households; Kibera: 92%, 23/25 households) and eggs (Dagoretti South: 88%, 22/25 households; Kibera: 68%, 17/25 households), or for income generation (Dagoretti South: 44%, 11/25 households; Kibera: 80%, 20/25 households) (Supplementary Table 4). Over half (56%; 28/50 households) of respondents reported allowing poultry and canines into the household (Supplementary Table 4). Study staff observed less animal feces near household soil sampling areas in Dagoretti South (24%; 6/25 households) than in Kibera (48%; 12/25 households).

### Strain identification of colonies by sample type

A total of 321 samples (132 human stool, 111 poultry cloacal swabs, 17 canine feces, 22 stored drinking water, and 39 household soil) were successfully collected at the 50 enrolled households, and from each sample, up to 5 isolates were pooled togser for PIC-seq. A total of 1,516 colonies were included. Each sample pool was sequenced and analyzed with StrainGE^22^ to estimate the number of strains present in each sample, and their corresponding representative genomes, using a database of 2,496 *Escherichia* and *Shigella* reference genomes, clustered at 99% k-mer similarity (Methods). We identified 800 total strains that corresponded to 299 unique representative reference strains (Fig. 1A). The identified reference strains encompassed 279 *Escherichia coli*, 1 *Escherichia albertii*, 6 *Escherichia fergusonii*, and 13 strains within the genus *Shigella* (Fig. 1B). Out of the 13 identified *Shigella* strains, 11 were assigned to *Shigella flexneri*, and the remaining two were assigned to *Shigella dysenteriae* and *Shigella* sp.

**Fig. 1.**
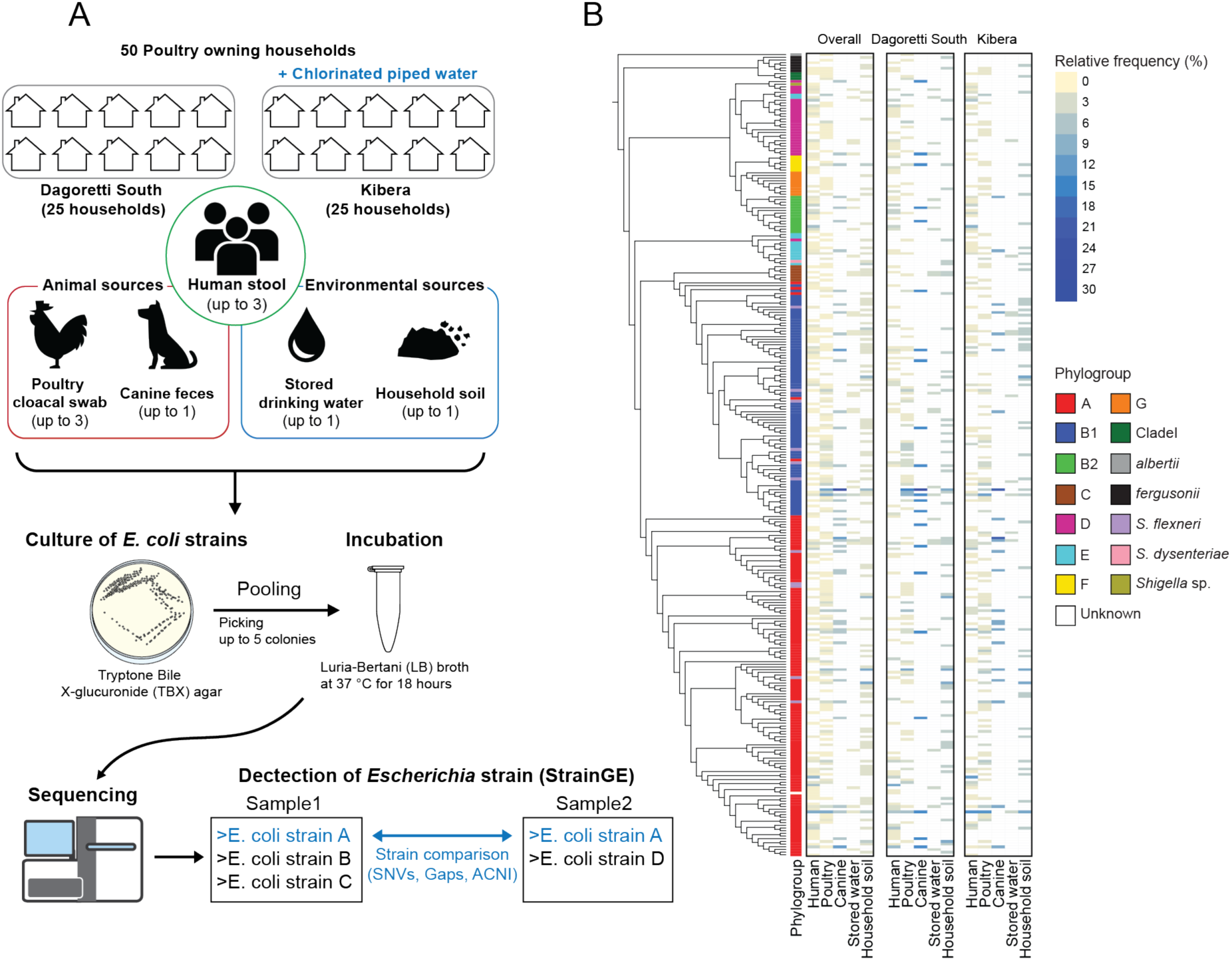
(A) Schematic of sample collection and pooling *E. coli* isolates cultured for sequencing. (B) Cladogram of all isolates built using maximum likelihood based on the alignment of core genes from genomes of reference strains with 1000 bootstraps. The heatmap shows relative frequency of reference strain detection in each sample type, either from overall households or households within each subcounty.

The number of identified strains in each sample was compared to assess sample strain diversity across sample types. Stored drinking water had significantly lower counts of strains compared to other sample types, and household soil had significantly higher strain counts than human stool (Supplementary Fig. 1A); no other significant differences were observed. We then investigated how distinctively strains are distributed within sample types, based on their representative reference strains. To account for different sample sizes of each sample type, we subsampled an equal number (*n* = 10) of samples with 1,000 iterations and compared the number of unique reference strains (Supplementary Fig. 1B). Of all sample types, household soil samples exhibited the highest number of unique reference strains while stored drinking water samples exhibited the lowest. The number of unique reference strains from poultry cloacal swabs and canine feces was greater than from human stool samples. In human stool, the number of unique reference strains increased with age (Supplementary Fig. 1B).

We compared the phylogroup compositions of the representative reference *E. coli* strains identified across sample types. Phylogroup A was the most abundant with 31.4% to 46.4% of all identified strains across all sample types (Supplementary Fig. 2). The next most common phylogroup was B1, although its frequency among strains identified from humans was noticeably lower (17.2%) than in other sample types (30.4% – 37.1%). Phylogroups B2 and D were found to be enriched in humans (phylogroup B2: 12.3%; phylogroup D: 11.7%), stored drinking water (phylogroup B2: 8.6%; phylogroup D: 11.4%), and B2 in canine samples (6.3%), compared to other sample types (phylogroup B2: 0.3% – 0.9%; phylogroup D: 3.4% – 4.8%). Additionally, *S. flexneri* was more frequently found among the strains from poultry (7.5%) and canine (4.2%) samples, compared to other sample types, which ranged from 0% to 2.6%.

### Prevalence of *E. coli* pathotypes and *Shigella*

To examine the prevalence of *E. coli* pathotypes across sample types, we investigated the presence of known virulence genes on the assembled contigs for each sample (Methods). Humans carried the most diverse pathotypes of *E. coli*, with Enteroaggregative *E. coli* (EAEC) (13.6% of human samples) and Enteropathogenic *E. coli* (EPEC) (7.6%) most frequently detected (Fig. 2). EAEC and EPEC were also frequently detected in canine feces (EAEC: 5.9% and EPEC: 5.9% of canine samples). In poultry cloacal swabs, EAEC (1.8% of poultry samples), EPEC (0.9%), and Enterotoxigenic *E. coli* (ETEC) (0.9%) were detected. In stored drinking water, EAEC (4.1% of drinking water samples) was the only pathotype detected while household soil had more diverse pathotypes present including EAEC (7.7% of soil samples), ETEC (7.7%), and EPEC (2.6%). Diffusely adherent *E. coli* (DAEC) and Enteroinvasive *E. coli* (EIEC) were not detected in any of the samples. We also determined the prevalence of *Shigella* across samples, which was highest in poultry cloacal swabs (19.0% of poultry samples), followed by canine feces (11.8% of canine samples), household soil (10.3% of soil samples), and human stool (6.1% of human samples) (Fig. 2). No *Shigella* strains were detected in stored drinking water.

**Fig. 2.**
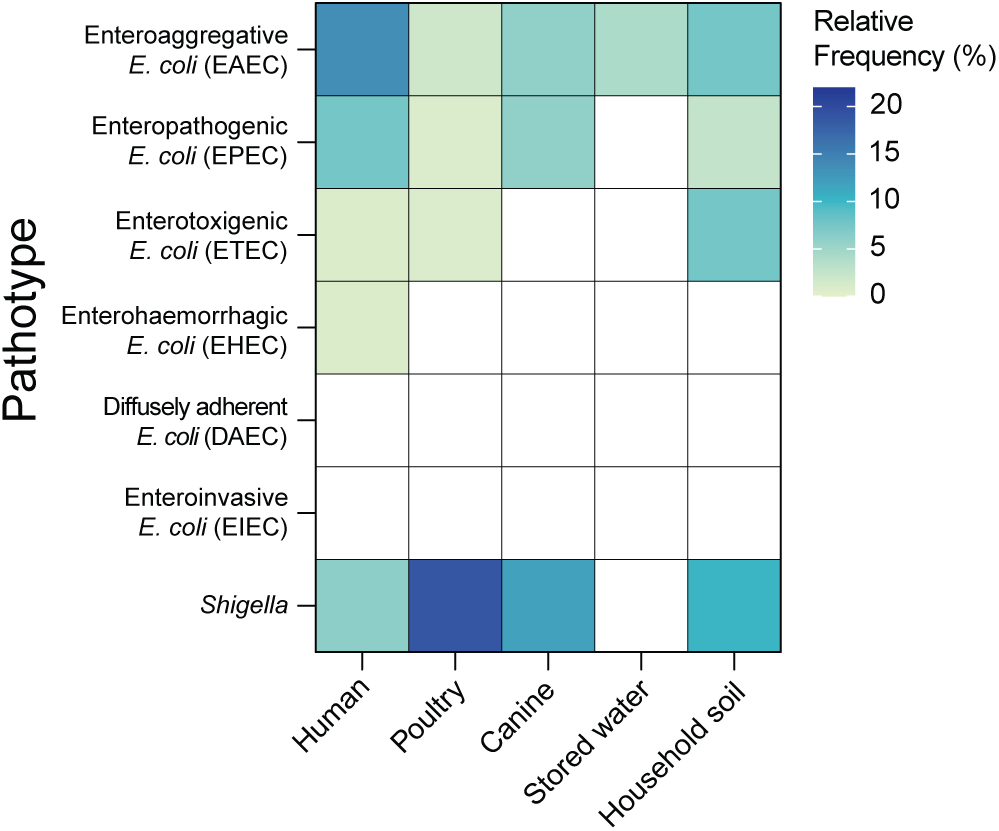
Prevalence of *E. coli* pathotypes and *Shigella* strains across sample types. Detection of *E. coli* pathotypes based on specific sets of virulence genes associated with each pathotype. The presence of *Shigella* was identified by StrainGE.

### Strain-sharing patterns

We examined strain-sharing patterns across humans, poultry, canine, soil, and stored drinking water *within* and *between* households. Sample pairs with strains with over 99.95% average callable nucleotide identity (ACNI) by StrainGE were considered to share strains based on previous benchmarking,^22^ and the average strain-sharing rate (the average strain-sharing events/total possible pairs in each household or household pair) *within* or *between* households were compared across different sample type pairs. *Within* households, strain-sharing was most frequent between the same host type (human-human rate: 0.19 cases per sample pair; poultry-poultry rate: 0.15 per sample pair) with rates significantly higher than the rates *between* households (human-human rate: 0.0051, *p* < 0.001; poultry-poultry rate: 0.0023, *p* < 0.001) (Fig. 3). Direct human-animal strain-sharing was nearly absent; a strain shared between human and poultry was detected in just one household, with the rates observed *between* households not significantly different from *within* households. Strain-sharing with environmental sources (human-stored water rate: 0.065; poultry-soil rate: 0.038) was frequently observed *within* households and at significantly higher rates than *between* households (human-stored water rate: 0.00030, *p* < 0.001; poultry-soil rate: 0.0024, *p* = 0.0048). Additionally, the strain-sharing rate between stored drinking water and soil *within* households (rate: 0.053) was significantly higher than *between* households, where no strain-sharing was detected (*p* < 0.001). Unlike *within* households, where strain-sharing was most common between human and stored drinking water, significantly higher than that between human and animal (*p* = 0.027), human-stored drinking water strain-sharing was one of the lowest *between* households (rate: 0.00030) (Fig. 3 and Supplementary table 5).

**Fig. 3.**
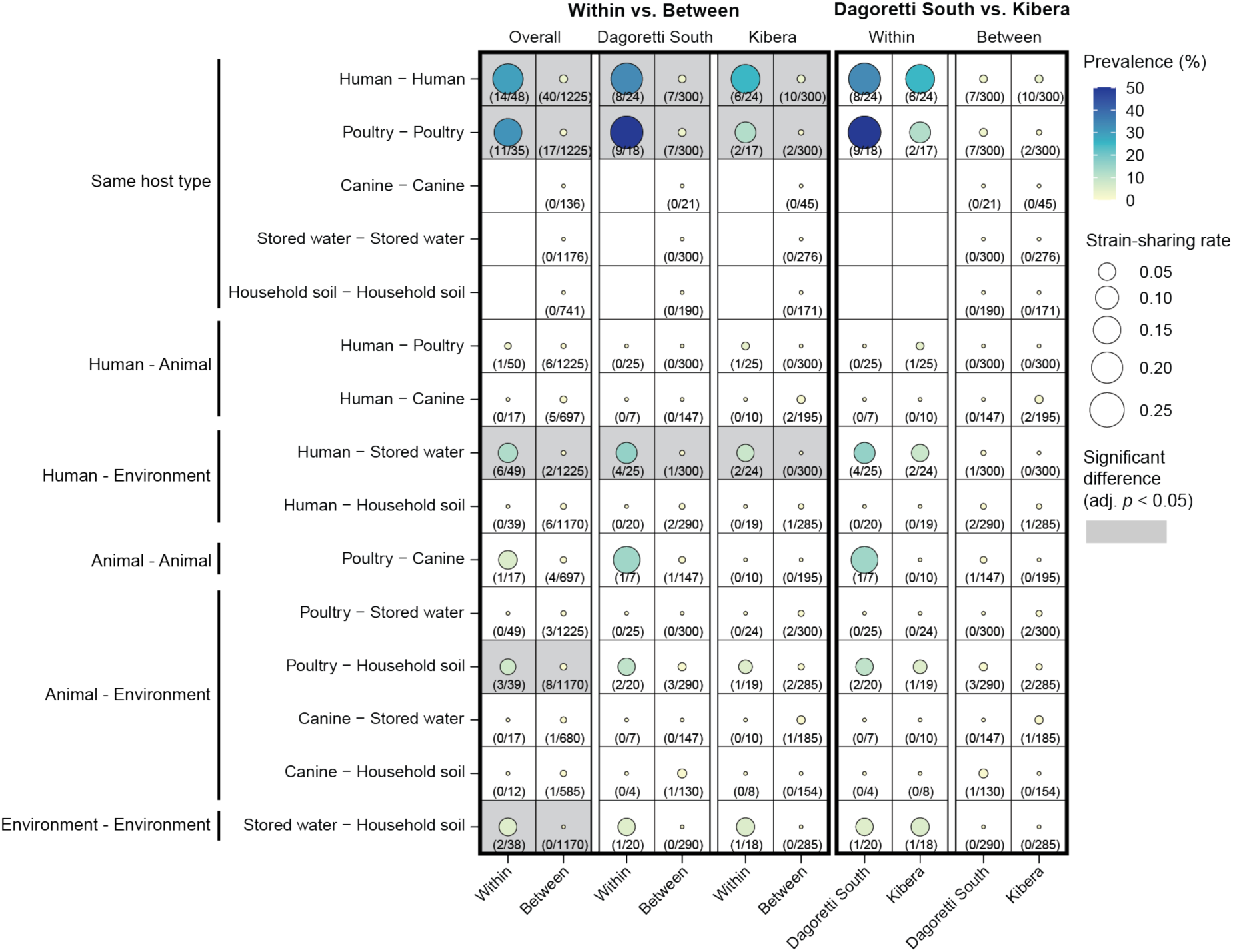
Strain-sharing patterns within and between households. The size of each circle corresponds to the strain-sharing rate (average strain-sharing events/total possible pairs in each household or household pair). The color gradient of each circle represents the prevalence of strain-sharing, calculated as the percentage of households or pairs of households where strain-sharing was detected relative to the total number of households or household pairs, with actual numbers provided as texts below each circle. Permutation tests with 1,000 bootstraps were used for testing for significant differences between mean strain-sharing rates, and all resulting *p*-values were adjusted using Benjamini-Hochberg correction.

### Contaminated drinking water facilitates human-human strain sharing

In Kibera, households had access to piped chlorinated drinking water. We observed poorer water quality in Dagoretti South; 52.0% of households (13 out of 25) in Dagoretti South had *E. coli* contamination in stored drinking water while 37.5% of households had *E. coli-*contaminated water in Kibera (9 out of 24) (Fig. 4A). We hypothesized that chlorinated water could reduce human-drinking water strain-sharing *within* households. Indeed, we observed less human-drinking water strain-sharing in Kibera (rate: 0.049) than Dagoretti South (rate: 0.080) although the difference was not statistically significant (Fig. 3). Given that humans *within* households share the same drinking water source, contaminated stored drinking water may play a role in strain-sharing between humans *within* households. Indeed, human-human strain-sharing was observed in 42.9% (9/21) of the households with *E. coli*-contaminated stored drinking water, compared to just 19.2% (5/26) of the households without *E. coli* contaminated water (Fig. 4B). The human-human strain sharing (rate: 0.32) *within* households with *E. coli*-contaminated stored water was significantly higher than households with clean water (rate: 0.090, *p* = 0.026) (Fig. 4B). No other strain-sharing sample type pairings showed significant differences when households were stratified by stored water contamination (data not shown). Further, we observed identical strains being shared across more than one human and stored drinking water in two households (Household ID: 051 and 084), lending evidence for strain-sharing between humans through the consumption of contaminated stored drinking water (Fig. 4C).

**Fig. 4.**
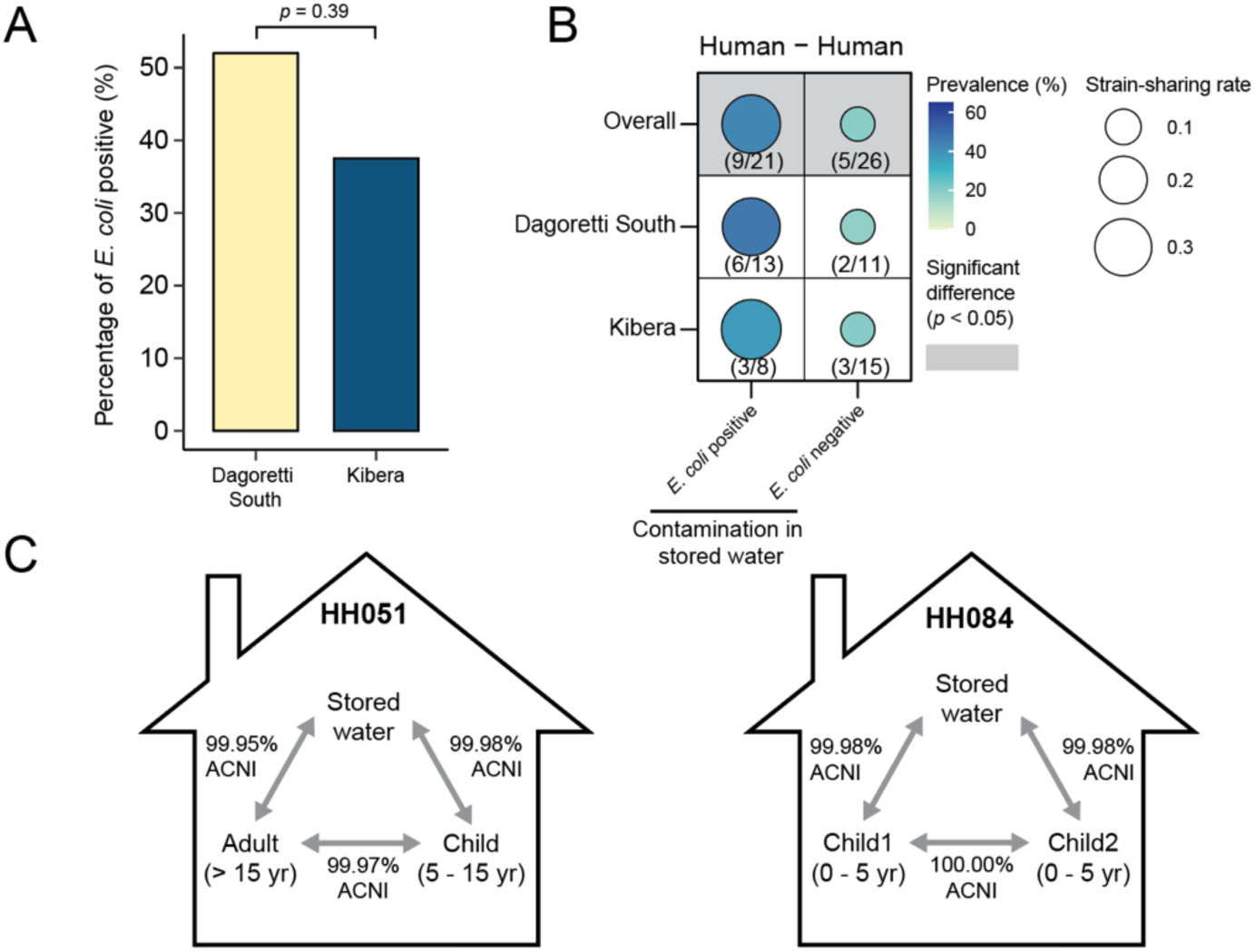
Strain sharing within households stratified by presence of *E. coli* contamination in stored drinking water. (A) Proportion of households with *E. coli* contaminated stored water in Dagoretti South and Kibera. (B) Comparison of human-human strain-sharing within households with and without *E. coli* contamination in stored drinking water. (C) Observation of strain-sharing patterns in two households with the same strain shared across humans and stored drinking water within households.

### PIC-seq outperforms metagenomics

We additionally evaluated the performance of PIC-seq in investigating *E. coli* strain-sharing patterns by comparing the results with metagenome sequence data generated without the culture enrichment step. As part of another study, total DNA was extracted from a subset of matched human stool samples in this study (110 samples) and sequenced. StrainGE detected a slightly lower number of strains across the total metagenome samples (230 strains) vs. the corresponding PIC-seq samples (248 strains). Further, while the PIC-Seq approach detected strain-sharing in 50 sample pairs, strain-sharing was detected in only 7 metagenomic sample pairs (5 pairs overlapping between the two approaches), possibly due to insufficient coverage depth for detecting genomic substitutes in *E. coli* between samples (Supplementary Fig. 3).

### Distribution of clinically relevant ARGs across humans, animals, and the environment

To investigate ARG transmission, we examined the distribution and diversity of ARG variants based on the assembled contigs for each sample type. To assess if long-read sequencing provided higher resolution insights into ARG transmission, we benchmarked using simulated pooled *E. coli* datasets on contigs assembled from Oxford Nanopore Technology long reads versus Illumina short reads (Supplementary methods). While contigs assembled from long read data consistently generated longer contigs than those from short reads across all simulated datasets – in terms of the proportion of the recovered genome fraction, the length of the largest alignment, and N50 – they exhibited the lowest accuracy in their sequence identity to the original genomes (Supplementary Fig. 4). Hybrid assembly utilizing both short and long reads marginally improved contig length but resulted in lower accuracy compared to short-read assemblies. Thus, we opted to analyze ARGs using only contigs assembled from short reads. Additionally, we benchmarked our assembly approach, which uses reference-based binning before *de novo* assembly to reconstruct resistomes, against sole *de novo* assembly (Supplementary Methods). Our approach significantly outperformed sole *de novo* assembly, reconstructing 81.6% to 89.2% of the original resistome compared to just 15.1% to 27.8% (Supplementary Fig. 5).

We grouped all identified ARGs on the contigs into clusters based on their nucleotide sequences at 100% identity and compared the number of unique ARG clusters observed in each sample type. Similar to strain diversity patterns, household soil had the highest number of unique ARG clusters (median: 266) and stored drinking water the lowest (median: 152) (Supplementary Fig. 6). No significant difference was observed between humans (median: 200.5) and animals (poultry median: 231; canine median: 238).

We investigated the diversity and distribution of high-risk ARGs, classified as Rank I ARGs by Zhang *et al.*^30^, across our sample types. Rank I ARGs refer to gene families that have been reported to be significantly enriched in human-associated environments, associated with gene mobility, and are known to be present in ESKAPE pathogens (*Enterococcus faecium*, *Staphylococcus aureus*, *Klebsiella pneumoniae*, *Acinetobacter baumannii*, *Pseudomonas aeruginosa*, and *Enterobacter* sp.). Humans had significantly higher numbers of Rank I ARG clusters conferring resistance to the drug classes of beta-lactam and macrolide-lincosamide-streptogramin (MLS) than poultry, while Rank I ARG clusters of trimethoprim and quinolone were found to be higher in poultry (Supplementary Fig. 7). We assessed ARG mobility in our samples based on adjacency to mobile genetic elements (MGEs); if ARGs were associated with contigs under 5000 bp, they were determined to have ambiguous mobility (Methods). For ARGs where mobility could be determined, an average of 64.2% of ARGs within Rank I ARG clusters were predicted to be mobile, except for *mdtE*, where none of ARGs within the clusters were predicted to be mobile (Fig. 5 and Supplementary Table 6). Prevalence of ARG clusters across humans, animals, and the environment was similar, with several ARG clusters prevalent in all sample types. *aph(6)-Id*, which confers resistance to the antibiotic class of aminoglycoside, had the largest cluster size and was the most frequently present in all groups, including in 41.2% of canine samples. Among β-lactamase genes, a *bla*_TEM-1_ cluster was the most prevalent in all groups, with detection in 51.4% of poultry samples. Another β-lactamase gene cluster found in all sample types was *bla*_CTX-M-15_, which confers resistance to third generation cephalosporins (Fig. 5). Its prevalence was comparable in humans (20.5%), canines (23.5%), and household soil (15.8%); it had lower prevalence in poultry (4.5%) and stored water (5.3%). We identified several other ARG clusters in all sample types; genes conferring resistance to trimethoprim (*dfrA*), MLS (*mphA*), and quinolone (*qnrS*) were prominent. We also observed widespread presence across all samples of the bacitracin resistance gene *bacA*, a peptide antibiotic, and the *tolC* gene, associated with multidrug resistance as part of the efflux pump system.

**Fig. 5.**
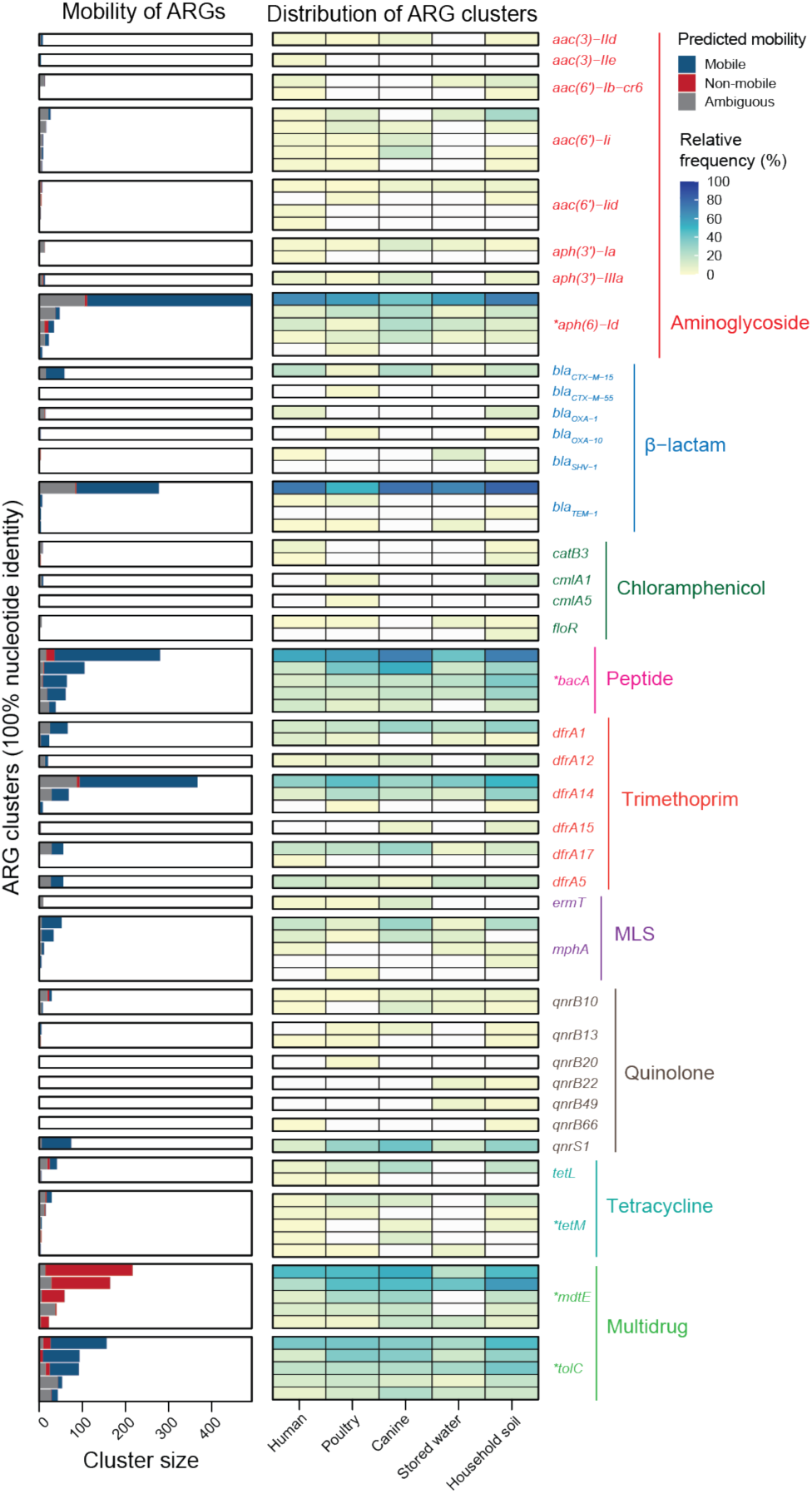
Distribution and mobility of clinically relevant ARG clusters at high-risk (Rank I) identified across humans, animals, and the environment. The asterisks on ARG annotations indicate that only the top five most abundant ARG clusters are depicted in the figure.

### Strain-sharing disseminates ARGs

To investigate whether spatial proximity is associated with similarity in ARG profiles, sample resistomes were compared *within* versus *between* households. Resistome similarity closely mirrored strain-sharing rates, with significantly higher resistome similarity *within* households compared to *between* households for human-human, poultry-poultry, human-water, and water-soil (Supplementary Fig. 8). In addition, poultry-canine pairs shared significantly higher resistome similarity *within* households than *between* household when stratified by predicted mobility (Supplementary Fig. 8).

To further investigate the influence of strain-sharing on resistome composition, we performed a correlation analysis between mean strain-sharing rates and mean resistome similarities between sample types (Fig. 6). We hypothesized that if ARGs are not specific to certain strains due to frequent horizontal gene transfer and are already prevalent among samples, a weak correlation between strain-sharing rates and resistome similarities would be expected. Both non-mobile and mobile resistome similarities were strongly correlated with strain-sharing rates *within* households (non-mobile vs. rate: Spearman’s *r* = 0.86; *p* = 0.0003 and mobile vs. rate: Spearman’s *r* = 0.89; *p* =0.000097), suggesting frequent strain-sharing *within* households was a strong factor for sharing similar resistomes (Fig. 6). However, no significant correlation was observed *between* households, presumably due to the insufficient number of strain-sharing events to affect mean resistome similarities. A bootstrapped analysis revealed different correlation coefficient distributions *within* versus *between* households (Supplementary Fig. 9).

**Fig. 6.**
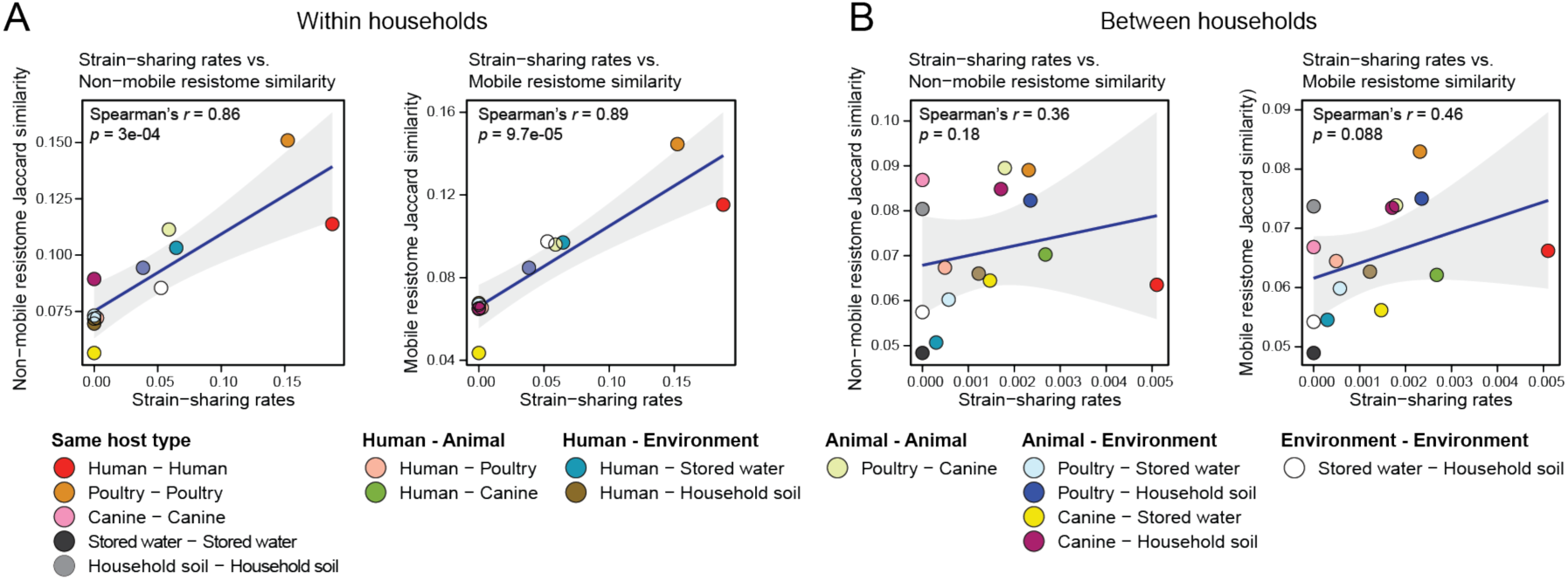
Spearman’s correlation analyses of non-mobile and mobile resistome similarity patterns with strain-sharing patterns. Left and right panels are the analyses (A) *within* and (B) *between* households.

## Discussion

Here, we observed high rates of *E. coli* strain sharing between humans and stored drinking water *within* households compared to *between* households, suggesting the importance of drinking water as a transmission route of bacterial strains to humans.^31,32^ *E. coli* contaminated drinking water was associated with higher human-human strain-sharing rates in households and we captured instances of the same strain shared between humans and water in the same household. Frequent strain exchange between drinking water and humans could explain the similarities in phylogroup profiles of strains between humans and stored drinking water, including the comparable proportion of phylogroups B2 and D.^33^ Our results are consistent with bacterial strains ingested through water consumption readily colonizing the human gastrointestinal tract.^34,35^ We observed lower levels of human-water strain-sharing rates and stored drinking water *E. coli* contamination in Kibera compared to Dagoretti South (although not statistically significant), suggesting access to chlorinated water in Kibera disrupted strain-sharing via drinking water.^36^ Water quality data collected by the parent study documented significantly lower levels of stored drinking water *E. coli* contamination in Kibera (22.6%)^37^ compared to Dagoretti South (68.2%) with a larger set of households (120 households) (Fisher’s exact test; *p* value < 0.001).^37^ A previous randomized controlled trial of community-wide passive chlorination of drinking water in urban Bangladesh found a 23% reduction in 7-day child diarrhea prevalence, suggesting the effectiveness of chlorinated water in disrupting bacterial transmission.^36^ Considering the complex bacterial transmission routes in densely populated urban informal settings,^8^ our findings highlight chlorinated drinking water as a key strategy to disrupt household strain-sharing.

It has been hypothesized that domestic animal exposure increases the risk of zoonosis.^31,38^ While we rarely observed direct zoonotic sharing events in our study population, we documented that poultry-soil stain sharing was more frequent *within* households compared to *between* households. In our urban study context, rare poultry-human strain sharing within households could be explained by infrequent exchange of strains between humans and soil (Fig 3). Previous research in rural and/or urban regions of Kenya,^39^ Bangladesh,^31,40^ and India^41^ has indicated that exposure to domesticated animals increases environmental fecal bacteria contamination, evidenced by the higher prevalence of animal versus human fecal markers in soil and drinking water. Given that environment-associated strain-sharing rates with humans or animals were significantly higher *within* households versus *between* households, our findings highlight the critical role of the environment in bacterial transmission. A few human-animal strain-sharing events were observed *between* households at a rate not significantly different from that *within* households, consistent with a previous study in Kenya, which found that sharing between humans and livestock was not confined to the same households.^13^ Taken together with previous evidence, our results suggest that human-animal strain-sharing can occur at the community scale.

Our results are consistent with clonal spread of bacteria being the main driver shaping resistomes of pooled *E. coli* strains. ARG variants associated with MGEs have the potential for horizontal gene transfer, which would result in overlapping ARGs between non-identical or phylogenetically distant strains. In our study, the significant correlation of mobile (and non-mobile) ARG similarity with strain-sharing rates between samples *within* households, indicates that close proximity strain-sharing had a greater influence on resistome similarities than ARG exchange via horizontal gene transfer. This observation aligns with a previous study in Kenya,^43^ where the multilocus sequence type of *E. coli* isolates from humans, domestic animals, and wild animals best explained the variance of the mobile ARG assemblages among them. Within households, human-water pairs showed higher resistome similarity (due to clonal transmission) than human-animal pairs, underscoring the role of frequent bacterial strain-sharing in ARG spread. Between households, where strain-sharing events were rare, no correlations were observed between resistome similarities and strain-sharing rates across sample types.

Environmental sample resistomes possessed many high-risk ARGs.^30^ Rank I ARG variants within environmental samples had similar or even higher diversity than human and animal hosts across antibiotics classes, and our results suggest new ARG variants can be transferred to humans and animals through drinking water and consumption of soil particles. ARGs like *bla*_CTX-M-15_, conferring resistance to third-generation cephalosporins, were detected consistently across all sample types. Use of antibiotics could then exert selection pressure in the gut enriching these ARGs. Quinolones and trimethoprim have been reported to be widely used by poultry farmers in Nairobi, Kenya, and this would explain the higher number of Rank I ARG clusters of these antibiotics observed in poultry compared to humans.^44,45^ Beta-lactam antibiotics have also been reported for use in animals in Nairobi;^44^ however, the observed higher diversity in humans could be attributed to frequent human consumption of these antibiotics.^46^ Prevalence of plasmid-mediated ARGs like *bla*_TEM-1_^47^ and *aph(6)-Id*^48^ across all sample types corroborated that certain ARGs and MGEs would be more prone to dissemination.^49^ Our profiling of ARG mobility was not always possible due to insufficient contiguity of partial genomic segments assembled from short reads. Oxford Nanopore sequencing technology, which generates longer sequences has the potential to identify the genomic context of ARGs.^50^ In our benchmarking, however, the contigs assembled with short reads in this study outperformed those with long reads in their sequence accuracy (Supplementary Fig. 4). Hybrid assembly, leveraging both short and long reads, has been suggested as an alternative approach with better assembly quality,^51^ however, the improvement in sequence contiguity and accuracy for hybrid assembly was marginal in our benchmarking.

Our results demonstrate the value of PIC-seq for examining strain sharing and the prevalence of ARGs. PIC-seq demonstrated higher performance in capturing more strain-sharing events (50 sharing events) compared to those identified through traditional metagenomic sequencing (only 7 sharing events) in a subset of matching stool metagenomes sequenced for another study (Supplementary Fig. 3). Further, our integration of additional *de novo* assembly with reference-based binning assembly enabled the recovery of ARGs located in genomic regions of pooled organisms that differed from reference genomes, outperforming sole *de novo* assembly (Supplementary Fig. 10). ARGs recovered from unbinned reads were mostly associated with MGEs, such as plasmids and genomic islands, which can circulate among strains via horizontal gene transfer.^52^ While these ARGs could not be tied to a specific strain, we were able to better capture the resistome of pooled isolates from each sample. Compared to sequencing of single isolates or sequencing the entire microbiome for strain tracking, our approach offers a distinct advantage in that it allows for simultaneous capture of strain identity and sensitive detection of ARGs with genomic context. PIC-seq is a scalable method for examining strain-sharing patterns that could be replicated for other bacterial species.

## Methods

### Study sites and sample collection

To investigate *E. coli* strain-sharing between humans, domesticated animals, and the environment, 50 poultry-owning households with at least one child under the age of 5 years were enrolled in two subcounties, Dagoretti South (n = 25) and Kibera (n = 25), in Nairobi, Kenya, during June – August 2019. One household was enrolled from each compound that owned poultry. Written informed consent was obtained from each adult participant prior to stool sampling and child assent and parental written consent were obtained for each child participant. A household survey was performed along with the sample collection to collect demographic information and household characteristics including water, sanitation, and hygiene access and practices.

Each household was visited twice for the sample collection. During the first visit, we collected up to 3 poultry cloacal swabs (depending on the number of poultry present), one canine fecal sample, one household soil, and one stored drinking water sample from each household. A trained veterinary student administered the poultry cloacal swabs and placed them in storage tubes filled with Cary-Blair transport medium, kept in a cooler for transport to the lab. To collect canine feces, the top layer of fresh feces from the center of the pile was transferred into a 50 mL centrifuge tube with a sterile plastic scoop. For household soil collection, a 30 cm x 50 cm square area closest to the entrance of the household was marked, and the top layer of the entire surface of the marked area was scraped with a sterile disposable plastic scoop. Approximately 150 g of soil was collected and transferred into a 500 mL sterile Whirlpak bag. The stored water within each household was collected by withdrawing 350 mL of the water from the bottom of the storage container with a sterile serological pipette. The water was then transferred into a 500 mL sterile Whirlpak bag containing 10 mg of sodium thiosulfate. All samples were immediately placed in a cooler filled with ice and transported to the Kenya Medical Research Institute (KEMRI).

Approximately 1 week from the initial visit, households were revisited to collect human stool from one household member in the following three age groups: child aged 0 – 4 years, child aged 5 – 14 years, and adult aged 15 years or older. A stool collection kit was provided during the first visit, which included a 50 mL plastic pot with a sterile scoop for each member with instructions on how to collect the sample. Caretakers were instructed to collect feces on aluminum foil, then, using sterile gloves and scoops, to transfer the feces to the plastic pot for collection. The primary caretaker of each household was informed by mobile phone one day prior to the revisit to collect stool from the previous night or the morning of the revisit day. If stool samples were not available on the visit day, households were revisited up to three times to collect the remaining stool samples. After pick-up, the received stool samples were placed in a cooler filled with ice and transported to KEMRI.

The study received ethical approval from the Kenya Medical Research Institute (KEMRI) Scientific and Ethics Review Unit (12/3823) and the Tufts Health Sciences Institutional Review Board (13205). Additionally, a research permit was granted by the Kenyan National Commission for Science, Technology, and Innovation.

### Pooling Isolated Colonies for sequencing (PIC-seq)

Here we describe our method of pooling presumptive isolated *E. coli* colonies for sequencing (PIC-seq). *E. coli* colonies were first cultured from the collected samples within 6 hours of sample collection. A sterile swab was inserted into 1 g of each collected human stool and canine feces sample, and the swab was streaked on Tryptone Bile X-glucuronide (TBX) agar plates. Poultry cloacal swabs were directly streaked on the agar plates. Household soil and stored water samples were membrane filtered and cultured on TBX. Collected soil was first screened through 2 mm-pore mesh sieves to remove rocks and leaves, then 5 g were homogenized with 50 mL of distilled water. After allowing soil particles to settle for 1 minute, 100 μL of supernatant was diluted with 10 mL of distilled water and vacuumed through a 0.45-μm membrane filter, and plated. 100 mL of stored water was directly filtered, then plated on TBX. All plates were incubated overnight (18 – 24 hours) at 44 °C.

Post incubation, up to 5 presumptive *E. coli* colonies (blue-green color) were selected from each agar plate for pooling. The selected colonies from each plate were transferred into a single 2 mL tube containing 1 mL of Luria-Bertani (LB) medium. The pooled *E. coli* colony cultures were incubated overnight (18 hours) at 37 °C with shaking at 200 rpm. After incubation, 100 μL of the cultures were mixed with 1 mL of 30 % glycerol-70% Tryptone Soy Broth (TSB) solution and stored at – 80 °C. The samples were then shipped on dry ice to Tuft University (Medford, MA).

Pooled isolates were thawed on ice and inoculated into 5 mL of fresh TSB medium with a sterile inoculating loop. The cultures were incubated overnight at 37 °C with 200 rpm shaking, then 1.5 mL of the culture was pelleted by centrifugation at 5,000 x g for 10 min. The DNA was then extracted from pellets using DNeasy PowerSoil Pro Kit (Qiagen, Hilden, Germany) following the protocol provided by the manufacturer. The concentration and purity of the extracted DNA were measured by a Qubit 4.0 Fluorometer (Thermo Fisher Scientific, Waltham, MA), a sequencing library was prepared using Nextera XT kit (Illumina, San Diego, CA), and shotgun sequencing was performed on Illumina NovaSeq 6000 (Illumina) with target sequencing depth of 1.5 Gb at the Broad Institute (Cambridge, MA).

### Identification of *E. coli* strains and calculation of household-wise strain-sharing rates

The quality of raw sequence reads was examined using FastQC,^53^ and low-quality reads were filtered out using Trimmomatic v0.39^54^ with parameters set to LEADING:3, TRAILING:3, SLIDINGWINDOW:4:15, and MINLEN:70. The subsequent reads were used for the identification and comparison of *Escherichia* strains using the software StrainGE v1.3.3, the software specifically designed for accurate calling and comparison of strains in the complex metagenome data based on k-mer comparison, bypassing assembly process.^22^ The database for StrainGE was constructed using the complete genome sequences (2,496 genomes) of the genus of *Escherichia* or *Shigella* downloaded from NCBI RefSeq database on August 20, 2022, by using the built-in database construction pipeline (‘ncbi-genome-download’, ‘kmerize’, ‘kmersim’, ‘cluster’, and ‘createdb’). The reference genomes in the database were clustered if they shared 99% of their k-mer content with a Jaccard similarity over 0.90, and to minimize redundancy, only the representative genome sequences of each cluster were retained, resulting in a final set of 885 genomes.

After strain identification, we utilized StrainGR, the second module of StrainGE, to call sequence variants among samples.^22^ Sequence reads were aligned to the genomes of the identified strains in each sample using bwa mem v0.7.17 with default parameters.^55^ For sample pairs with the same reference strain, the average callable nucleotide identity (ACNI) and gap similarity were calculated based on the callable regions, using ‘call’ and ‘compare’ command in StrainGR.^22^ The threshold for defining the same strain was set to > 99.95% ACNI.

The strain-sharing rates between sample types were calculated, using the results from StrainGR. Within each household, we normalized the number of sample pairs sharing the same strain by the total number of combinations between the certain sample types. We then computed the average rates based on the rates from all households for comparison. For strain-sharing rates between different households, we normalized the number of sample pairs with the same strain by the total number of possible sample pairs, between all combinations of different households. We then calculated the average rates from these inter-household rates for comparisons between certain types of samples. Stored water samples without *E. coli* contamination were included in calculating water-related total number of possible pairs.

### Phylogeny and phylogroup of detected *E. coli* strains

To infer the phylogenetic relationships among the strains identified from the samples, we utilized the reference genome sequences included in the StrainGE database. Prokka v1.14.5 was used to annotate genes, with the genus parameter set to *Escherichia*.^56^ The core genes that exist in 99% of all reference genomes were identified using Roary v3.13.0.^57^ The concatenated core gene alignment was generated using MAFFT, and SNP-sites v2.5.1^58^ was used to extract the alignment regions with single nucleotide polymorphisms (SNPs) to reduce the size of the alignment. The reduced alignment was uploaded to CIPRES Science Gateway v3.3^59^ (www.phylo.org), and a maximum likelihood tree was constructed using RAxML v8.2.12^60^ with the nucleotide substitution model set to GTRGAMMA with 500 bootstraps. *E. coli* phylogroup was predicted using ClermonTyping v20.03^61^ with default parameters.

### Annotations of antibiotic resistance genes and mobile genetic elements

The integrated assembly pipeline, which combines reference-based binning and *de novo* assembly, was used to generate contigs for ARGs and mobile genetic elements (MGEs) annotation (Supplementary Fig. 11). Plasmid sequences were assembled from sequencing reads using metaplasmidSPAdes v3.15.4^62^ with default parameters. Subsequently, the original reads were aligned and binned to the set of the assembled plasmid sequences and the reference genomes of strains identified through StrainGST, using mSWEEP v.2.0.0^27^ and mGEMs v.1.3.0^18^ with default parameters. For each reference strain, the reads binned to their genome sequences were individually assembled *de novo* using metaSPAdes v3.15.4^63^ with default parameters. The sequence reads that did not align to any sequences were separately assembled *de novo* using metaSPAdes. The gene-coding sequences in the assembled contigs were predicted using Prodigal v2.6.3^64^ with default parameters. The translated amino acid sequences of the predicted gene-coding sequences were annotated using DIAMOND blastp v2.0.14.152^65^ against the Comprehensive Antibiotic Resistance Database (CARD) v3.2.4^66^ with the parameters set to ‘--id 90’, ‘--min-orf 25’, and ‘--query-cover 80’. The annotations with the highest bitscore for each gene-coding sequence were retained for the downstream analysis.

To predict the mobility of ARGs, we examined the presence of any MGEs in the flanking regions of the annotated ARGs. We extracted up to 5000 base pairs from both upstream and downstream of ARGs on the contigs using a custom bash script. The mobileOG-pl pipeline was then used to annotate MGEs with mobileOG-db release beatrix-1.6 as the reference database,^67^ which is a manually curated database that includes sequences from eight different databases (ICEBerg, ACLAME, GutPhage Database, Prokaryotic viral orthologous groups, COMPASS, NCBI Plasmid RefSeq, immedb, and ISfinder), along with homologs of the manually curated sequences. ARGs with identified MGEs in their flanking region were presumed to be mobile, while those without MGEs were categorized as non-mobile. In instances where the contig length was insufficient (< 5000 base pairs) to detect MGEs in the flanking region, the mobility of these ARGs was considered ambiguous.

### Identification of pathogenic *Escherichia* and *Shigella* strains

The identification of pathogenic *E. coli* strains was inferred based on the presence of pathotype-associated virulence genes detected in the assembled contigs for each sample. These virulence genes were annotated from the contigs using ABRicate, in combination with the Virulence Factor Database (VFDB) (download in May, 2023).^68^ The presence of different *E. coli* pathotypes was based on the detection of the following gene sets: *agg*, *aat*, and *aai* for Enteroaggregative *E. coli* (EAEC); *eae* and *bfp* for Enteropathogenic *E. coli* (EPEC); *elt* and *est* for Enterotoxigenic *E. coli* (ETEC); *stx* for Enterohaemorrhagic *E. coli* (EHEC); *afa* and *drg* for Diffusely adherent *E. coli* (DAEC); and *ipa* for Enteroinvasive *E. coli* (EIEC).^69–71^ Presence of *Shigella* was additionally investigated based on StrainGST results.

## Statistical analysis

All statistical analyses in this study were carried out using R version 4.2.1.^72^ The differences in mean strain-sharing rates were compared using a nonparametric permutation test, with 1000 iterations of resampling. Resistome similarity was estimated by Jaccard similarity of ARG cluster compositions, defined as the number of unique ARG clusters shared divided by the total number of unique ARG clusters from both samples. The pairwise Mann-Whitney U test was used for comparing resistome similarities between sample types, using the ‘wilcox.test’ command from the ‘stats’ package in R.^72^ All *p*-values were adjusted accordingly using Benjamini-Hochberg correction. Finally, the correlation analyses among strain-sharing and resistome similarity patterns were performed using Spearman’s rank correlation method, as implemented in the ‘cor.test’ function from the ‘stats’ package in R.^72^

## Data availability

All raw sequence data were deposited in the NCBI database under BioProject accession number PRJNA1126668.

## Acknowledgments

This work was financially supported by the Bill and Melinda Gates Foundation (Grant number OPP1200651); the Tufts Clinical and Translational Science Institute (CTSI) Pilot Studies Program; a Tufts Center for Integrated Management of Antimicrobial Resistance (CIMAR) Pilot Award; and with federal funds from the National Institute of Allergy and Infectious Diseases, National Institutes of Health, Department of Health and Human Services, under grant number U19AI110818 to the Broad Institute. We thank Marieke H. Rosenbaum, Christine Imali, Anne Okello, Fredrick Okuku Owuor, Eric Kipchirchir Tanui, Glory Wairimu, Henry Emonje Mukabwa, Frank Odhiambo, Erica R. Fuhrmeister, Maya Nadimpalli, Tim Julian, Gretchen Walch, Deeksha Bathini, Gracie Hornsby, Jeremy Lowe, and the many KEMRI interns for their research assistance.

