## Supplemental Information for "Bacterial strain sharing between humans, animals, and the environment among urban households"

[4]Chan Zuckerberg Biohub – San Francisco

[5]Blum Center for Developing Economies, University of California, Berkeley, Berkeley, CA 94720

### Supplementary methods

#### Preparation of simulated datasets for benchmarking

To benchmark our bioinformatics pipeline, we generated simulated datasets that emulate sequence reads we would expect from PIC-seq on five *Escherichia coli* isolates. The paired-end Illumina sequence reads (2x150 bp) and the complete genome sequences of five clinically relevant antibiotic resistant *E. coli* isolates, possessing either $bla_{CTX\text{-}M\text{-}15}$ or *mcr-1* genes along with other antibiotic resistant genes (ARGs), were provided by Broad Institute. We characterized each isolate based on their complete genome sequences. The phylogroup of each *E. coli* isolate was predicted using ClermonTyping v20.03[1] with default parameters. MOB-suite v3.1.0 was used to identify any plasmid contents included in the genome files.[2] The gene coding sequences within genomes were predicted using Prodigal v2.6.3[3] and were annotated using DIAMOND blastp v2.0.14.152[4] against the Comprehensive Antibiotic Resistance Database (CARD) v3.2.4 with parameters set to minimum identity of 90%, minimum query cover 80%, and minimum amino acid length of 25. The proximity of mobile genetic elements (MGEs) to genes annotated as ARGs was examined in a 5000 bp region both upstream and downstream of the ARGs, using the mobileOG-pl pipeline and the mobileOG-db release beatrix-1.6 as the reference database.[5]

To generate simulated datasets, we subsampled the sequence reads from each *E. coli* isolate and pooled with 6 different proportions, ranging from equal ratios to 400-fold difference in sequence coverage, targeting a total sequencing depth of 2 Gb using seqtk v1.3 (Supplementary table 7). After pooling sequences, the quality of the reads was screened using Trimmomatic v0.39[6] with parameters set to LEADING: 3, TRAILING: 3, SLIDINGWINDOW: 4:15, and MINLEN: 70. The downstream analyses were performed subsequently based on these datasets.

#### Comparison of resistomes reconstructed from two different assembly strategies

To evaluate the efficacy of using a reference-based binning approach prior to de novo assembly in reconstructing resistomes, we compared resistome profiles generated from assemblies with and without the binning process. 1) For assemblies that employed the binning approach, the procedure followed was identical to the approach used for real samples. The plasmid-like contigs were first assembled out of initial data, and the initial reads were aligned to these plasmid contigs and the reference genomes of strains predicted by StrainGE[7] using mSWEEP v2.0.0[8] and mGEMs v1.3.1[9] with default parameters. The reads binned to each reference genome were individually *de novo* assembled using metaSPAdes v3.15.4.[10] The unaligned reads were separately *de novo* assembled using metaSPAdes. 2) The assembly without binning process was simply performed by running metaSPAdes on the initial simulated dataset with default parameters.

For all contigs assembled from two different approaches, the coding gene sequences were predicted using Prodigal v2.6.3,[3] and the genes were annotated by running DIAMOND blastp v2.0.14.152[4] against the Comprehensive Antibiotic Resistance Database with the same parameter used for the real samples. The co-localization of MGEs was investigated by annotating coding genes located within 5000 bp both upstream and downstream of identified ARGs. ARGs that had MGEs in their flanking regions were considered to have the potential for mobility. ARGs with no MGEs in their flanking regions, but whose flanking region were shorter than 5000 base pairs due to contig size limitations, were categorized as having ambiguous mobility. The coding regions of all ARGs were clustered at 100% nucleotide identity together with ARGs from reference genomes for the comparison of resistomes.

**Assembly using short or long sequence reads of simulated datasets**

To examine the efficacy of utilizing long read sequence data for investigating ARG sharing dynamics, we compared the assembly metrics of contigs assembled using short or long sequence reads. The long sequence reads of the matched five *E. coli* isolates were generated using Oxford Nanopore Technology MinION Platform and provided by Broad, along with the paired-end Illumina reads. We generated the simulated PIC-seq *E. coli* dataset for long reads using the methods described for the short reads.

In addition to the contigs assembled using only short reads, we employed two additional approaches; 1) assembling using short reads first, then correcting with long reads and 2) first assembling long reads and then polishing the resulting contigs with the short reads mapped onto them. Binning of sequence reads to the reference genomes identified by StrainGE was performed before assembly, as described for the short reads. To assemble contigs starting with short reads and then correct the resulting contigs with long reads, we used HybridSPAdes v3.15.4[11] with default parameters. For the assembly of contigs beginning with long reads, we utilized metaFlye v.2.8.1[12] with default parameters, followed by medaka v1.7.0 for error correction in the contigs, using the same long reads as input. Short reads were then mapped on to the contigs using bbmap,[13] and the resulting alignment files were utilized to polish errors in the contigs using pilon v1.24.[14] The assembly metrics of the contigs were estimated by using MetaQUAST v5.2.0,[15] based on reference genomes provided as the complete genomes of *E. coli* isolates used for simulated datasets.

### Supplementary results

#### Strain identification of the simulated PIC-seq datasets

The characteristics of *E. coli* strains used for generating the simulated PIC-seq data were analysed based on their complete genome sequences. Genome sizes across the strains ranged from 4.7 to 5.3 Mbp. The isolate GTEN 247 had the highest number of plasmids while no plasmid sequences were detected in the GTEN 378 isolate (Supplementary Table 8). In terms of phylogroup classification, the *E. coli* strains consisted of phylogroups commonly observed in real samples, with the isolates GTEN 291 and GTEN 293 assigned to identical phylogroup A (Supplementary Table 8). The range of unique ARG clusters identified across the strains was comparable, varying from 45 to 57. The GTEN 378 isolate had the lowest number ARGs predicted to be mobile, possibly due to the absence of plasmid. When individual strain identification was conducted on the Illumina sequences for each isolate, distinct strains were identified for each one.

Subsequent strain identification was conducted on simulated PIC-seq datasets generated by pooling sequence reads from *E. coli* strains in varying proportions. In all datasets, only four strains were identifiable, and the reference strain initially associated with GTEN 291, which was *E. coli* NCTC9087, was not identified, possibly due to high similarities (> 99.0% ANI) between GTEN 291 and GTEN 293 (Supplementary table 7). Except for GTEN 291, the trends of the relative abundance of the identified strains and the pooled ratio were generally in good agreement.

#### Benchmarking of reconstructed resistomes

The purpose of performing PIC-seq on *E. coli* isolates was not only to have a higher number of strains investigated for strain-sharing events but also to examine the resistomes with their genomic contexts. Sequence data comprising up to five different isolates is expected to yield better contig reconstruction compared to sequence data from more complex microbial communities. However, the assembly process could be challenging in deconvoluting pooled strains as the strains were highly similar. We employed the assembly approach suggested in plate sweep metagenomics, which involves a preliminary binning process based on reference strains before *de novo* assembly. In addition to assembly of binned reads, an additional *de novo* assembly was performed on the reads that did not map to any of the reference genomes, and ARGs and MGEs were annotated on the entire contigs. We performed benchmarking tests on this bioinformatics pipeline to evaluate its performance in reconstructing the resistome of samples, using simulated datasets. We also compared the resistomes reconstructed from contigs that were *de novo* assembled without a binning process to evaluate whether the binning process enhanced the accuracy of resistome reconstruction.

The assembly incorporating a binning process consistently outperformed those that did not in reconstructing resistomes across all simulated datasets. With a clustering at 100% nucleotide identity, the simulated dataset was expected to contain 212 overall ARG clusters, comprising 118 non-mobile and 94 mobile ARG clusters (Supplementary Fig. 5). In resistomes with a binning process, the number of overall true-positive ARG clusters, which aligned with those from reference genomes, ranged from 173 to 189 (accounting for 81.6% to 89.2%). In contrast, resistomes without a binning process only had 32 to 59 overall true-positive ARG clusters (accounting for 15.1% to 27.8%) (Supplementary Fig. 5). The number of false-positive overall ARG clusters, defined as those deviating from ARG sequences in the reference genomes, was comparable between resistomes reconstructed with and without a binning approach in the simulated datasets of ratio 1, 2, and 3. However, in the simulated datasets with ratios 4, 5, and 6, which had at least a 100-fold difference in the pooling ratios between strains, the number of false-positive ARG clusters was significantly lower in resistomes reconstructed without the binning process (fewer than 10 instances). The same trends were observed in both non-mobile and mobile resistomes. The non-mobile resistome had a higher number of true-positive ARG clusters, while the mobile resistome had a lower number of false-positive ARG clusters.

To determine which approach more accurately captures the true resistome, we calculated the Jaccard similarity between the resistomes derived from the simulated datasets and the reference genomes. In all the simulated datasets, resistomes reconstructed using the binning process exhibited at least a 1.9-fold higher Jaccard similarity to the reference compared to those without binning. This suggests that the incorporation of a binning process in the assembly pipeline significantly enhances the accuracy of resistome reconstruction in this study. (Supplementary Fig. 10).

**Supplementary figures**

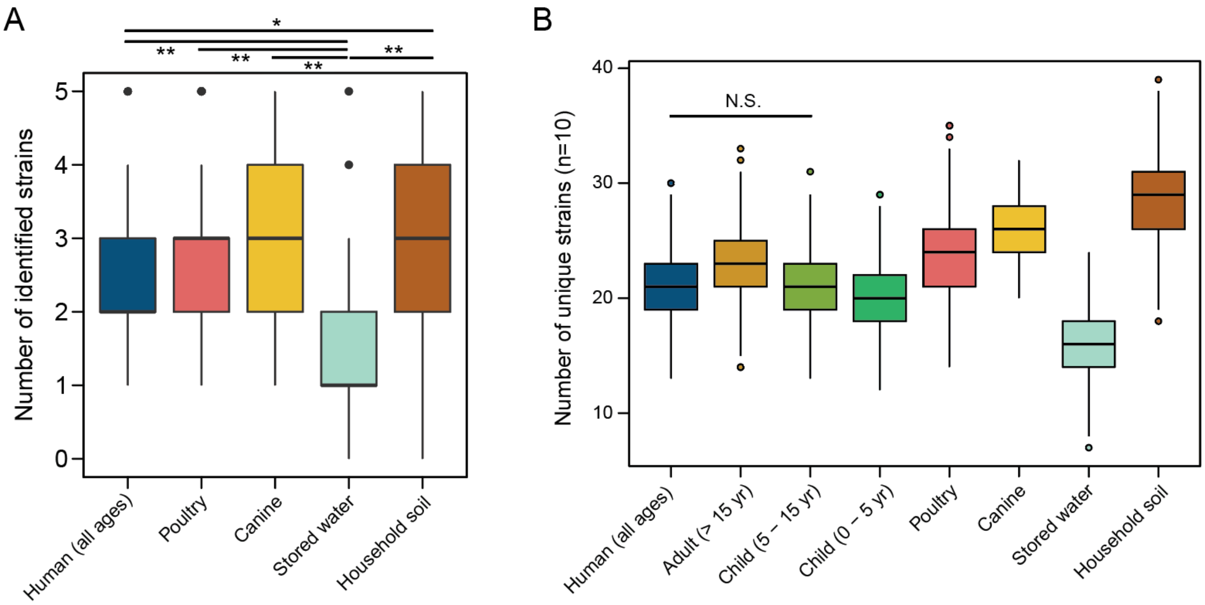


**Supplementary Fig. 1** (A) Number of strains identified by StrainGE in each sample across sample types. Bars marked with asterisks denote statistically significant differences (*: adjusted $p < 0.05$; **: adjusted $p < 0.01$). (B) The number of unique strains identified in subsampled sets ($n = 10$) through 1,000 iterations, categorized by host type. Bars marked with N.S. indicate no significant difference, while others show significant differences as determined by the Mann-Whitney U Test (adjusted $p < 0.05$).

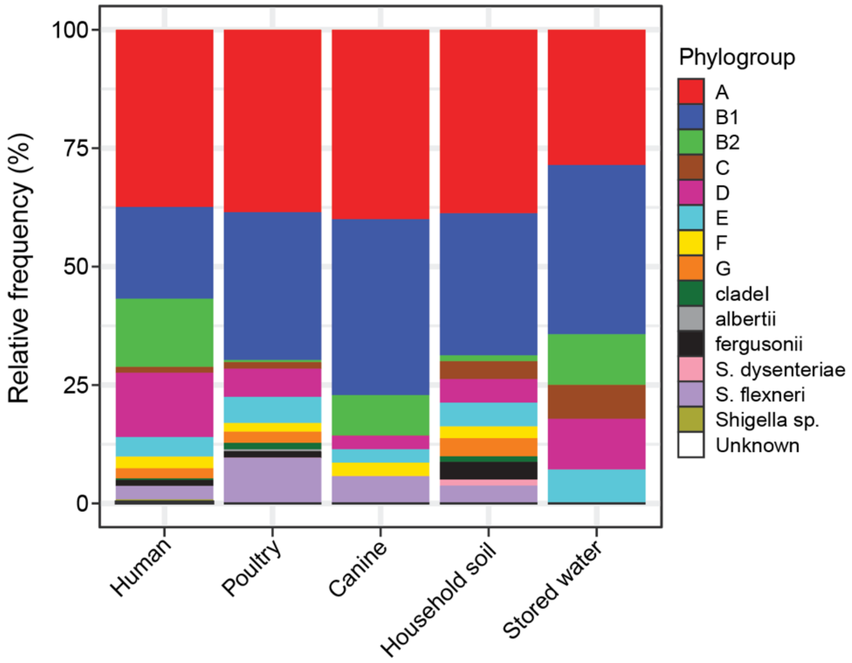


**Supplementary Fig. 2** Phylogroup composition of the identified representative reference strains by StrainGE, across sample types.

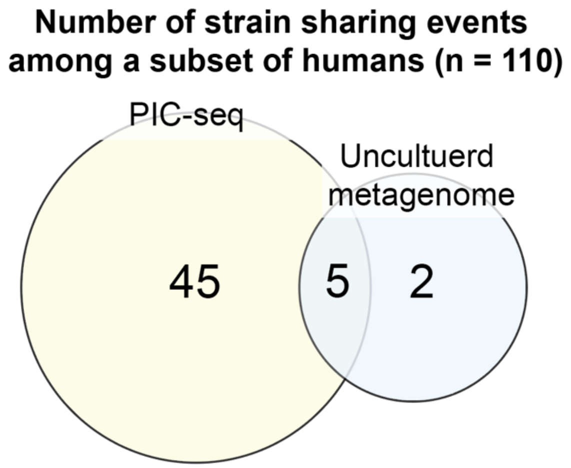


**Supplementary Fig. 3** Comparison of strain-sharing events between PIC-seq in this study and uncultured metagenome sequencing from a subset of matching samples (110 human stool samples).

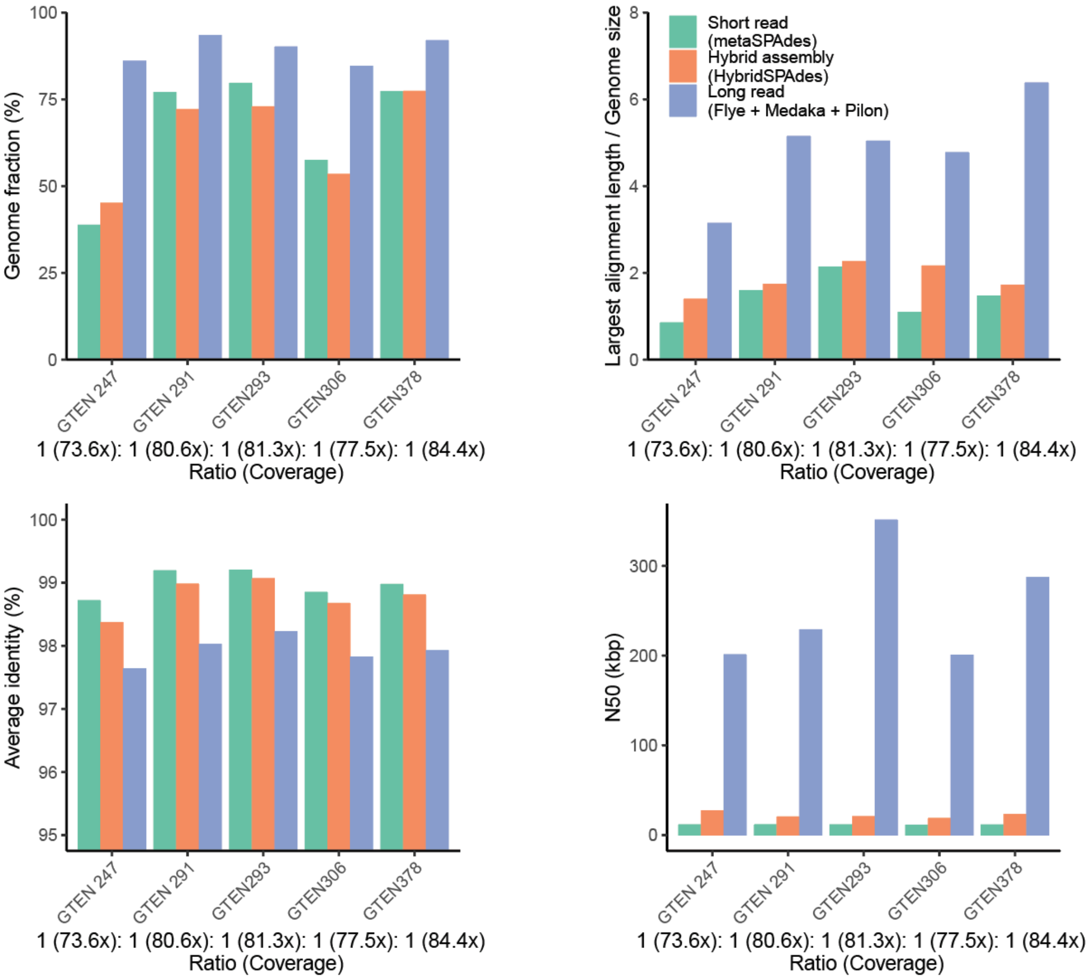


**Supplementary Fig. 4** Assembly metrics for contigs assembled using short or long sequence reads. The simulated PIC-seq dataset of *E. coli* strains with an equal ratio of reads was used. The numbers in parentheses on x-axis specify the actual sequence coverage of each genome. In the fill color legend: *Short read* indicates the contigs assembled using only short reads; *Hybrid assembly* indicates the contigs initially assembled with short reads, subsequently error-corrected using long reads; *Long read* indicates the contigs that were assembled first using long read, then error-corrected with short reads aligned onto them.

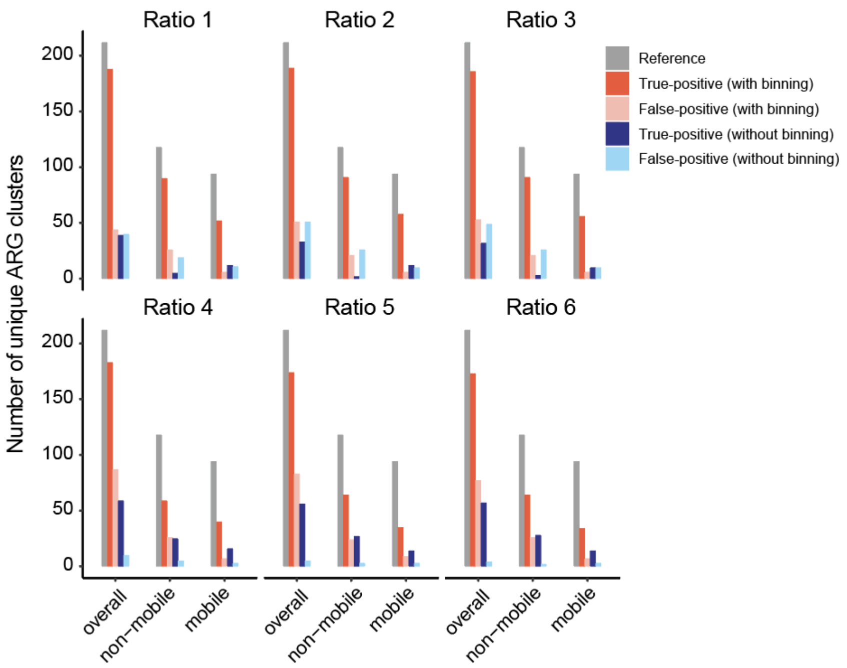


**Supplementary Fig. 5** Number of unique ARG clusters in the benchmarking test of the bioinformatics pipelines using the simulated datasets.

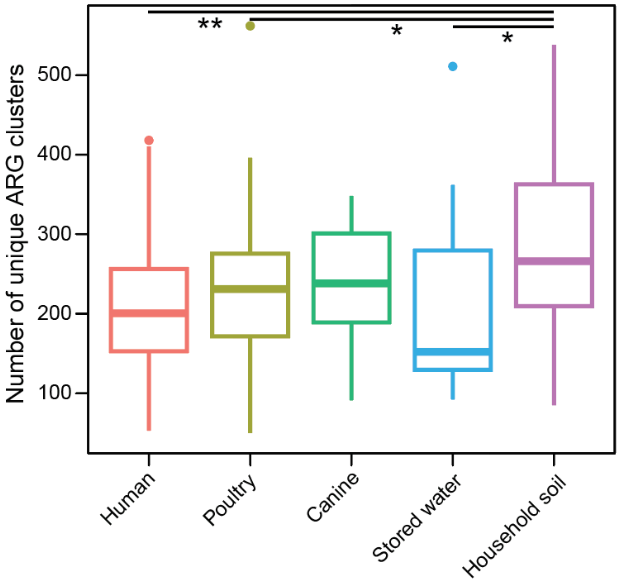


**Supplementary Fig. 6** Number of unique ARG clusters in each sample across sample types. Bars marked with asterisks denote statistically significant differences as determined by the Mann-Whitney U Test (*: adjusted $p < 0.05$; **: adjusted $p < 0.01$).

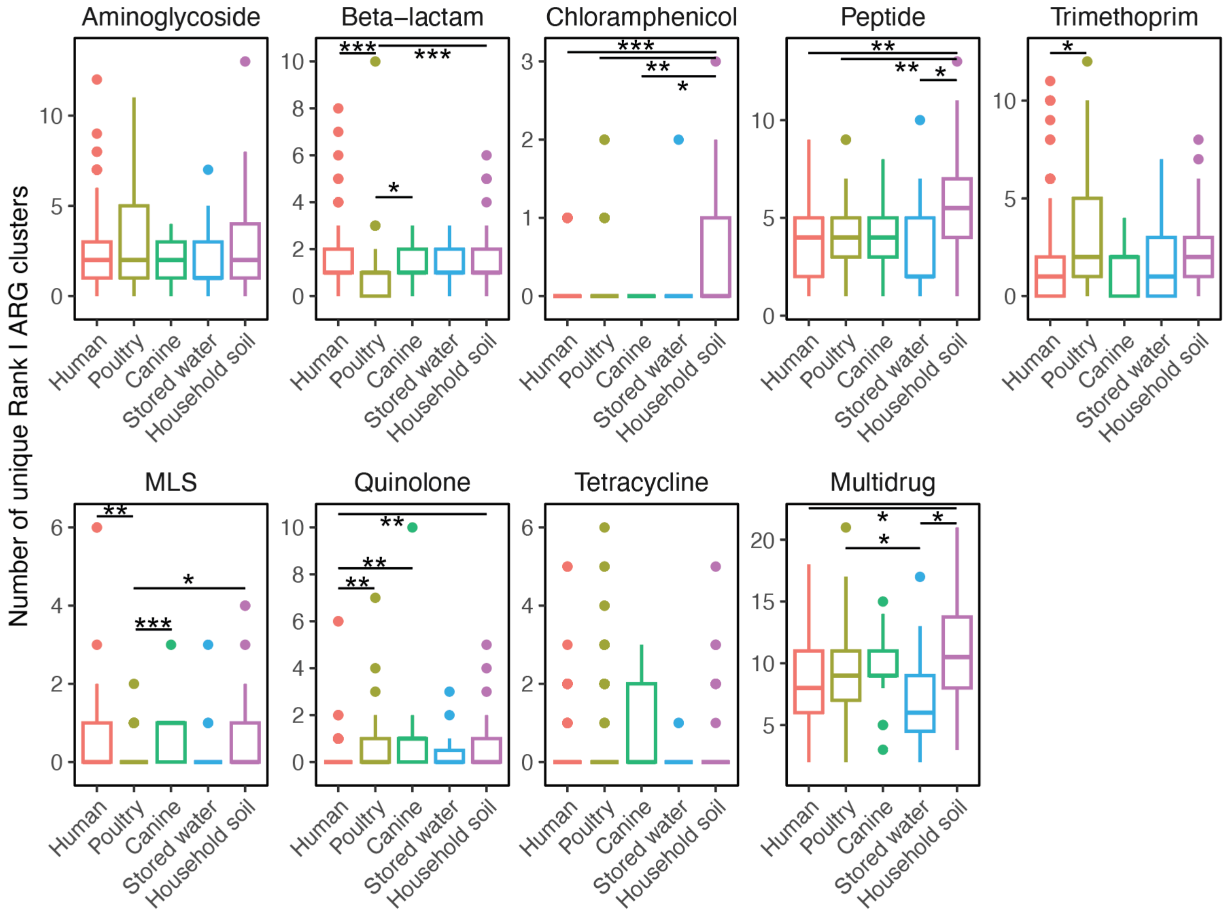


**Supplementary Fig. 7** Number of Rank I[29] unique ARG clusters in each sample across sample types grouped by drug classes. Bars marked with asterisks denote statistically significant differences (*: adjusted p < 0.05; **: adjusted p < 0.01; (***: adjusted p < 0.001).

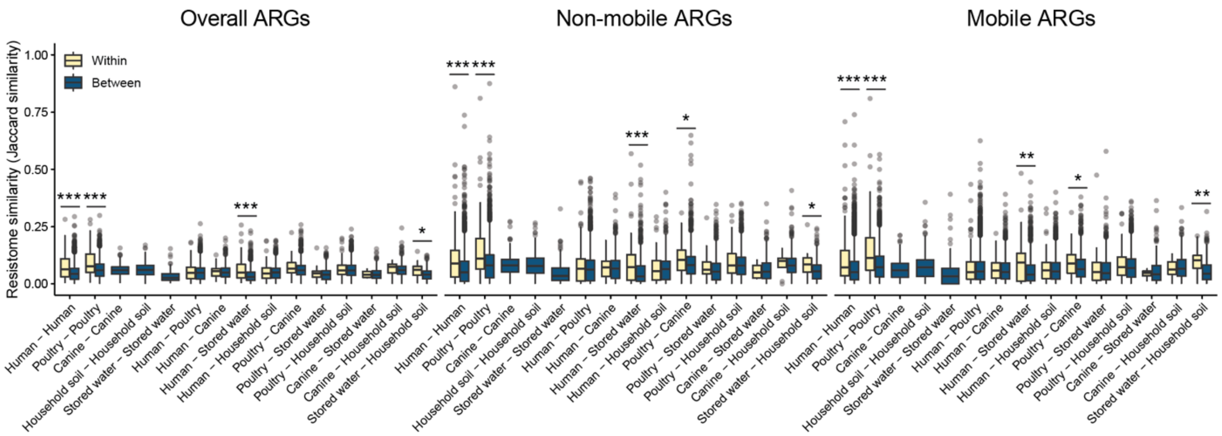


**Supplementary Fig. 8** Comparison of overall, non-mobile, and mobile resistome similarities *within* versus *between* households calculated based on Jaccard similarities. The asterisk denotes statistically significant differences (*: $p < 0.05$; **: $p < 0.01$; ***: $p < 0.001$)

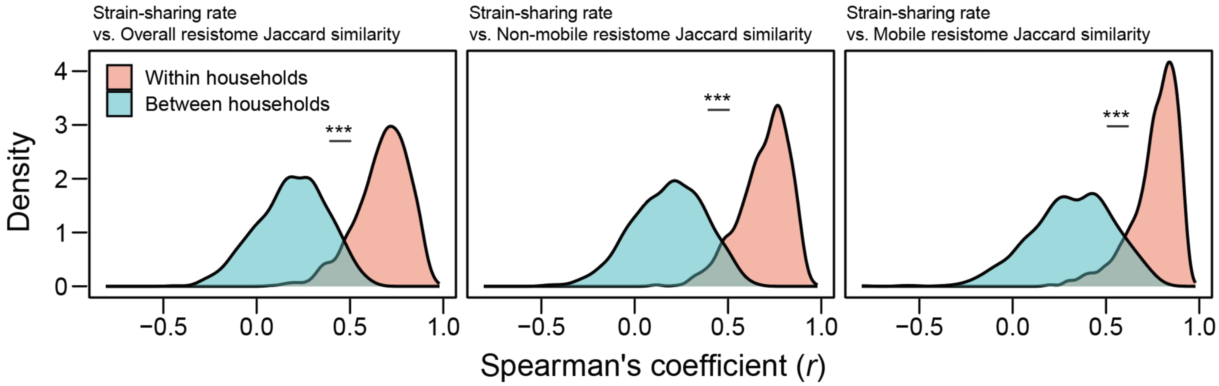


**Supplementary Fig. 9** Distribution of Spearman's coefficient (*r*) between resistome similarity and strain-sharing rate. The coefficients were calculated based on samples from resampled households bootstrapped 1000 iterations. The asterisks (***) indicate significant difference between the distributions (Mann-Whitney U test, $p < 0.001$)

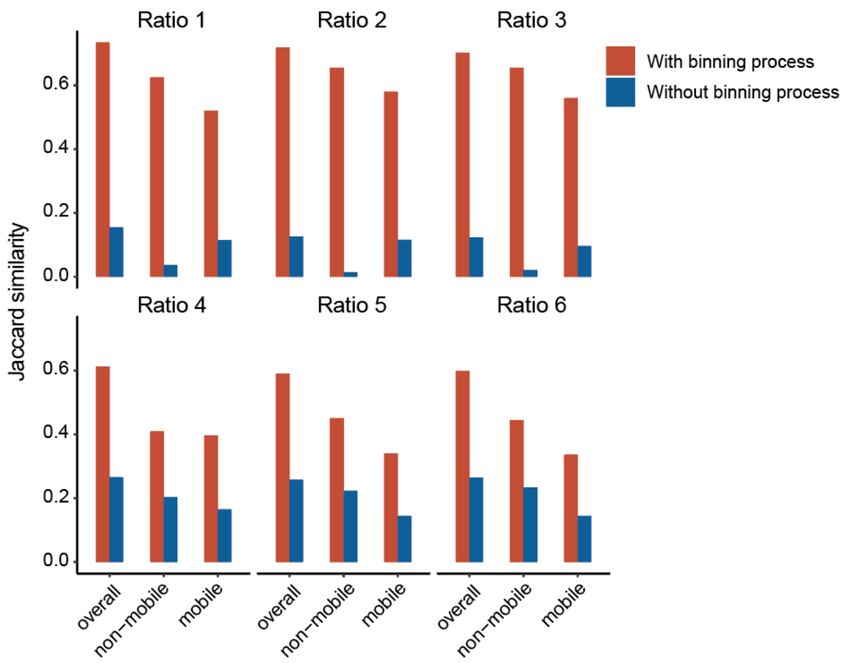


**Supplementary Fig. 10** Jaccard similarity of resistomes based on ARG clusters in benchmarking compared to the reference.

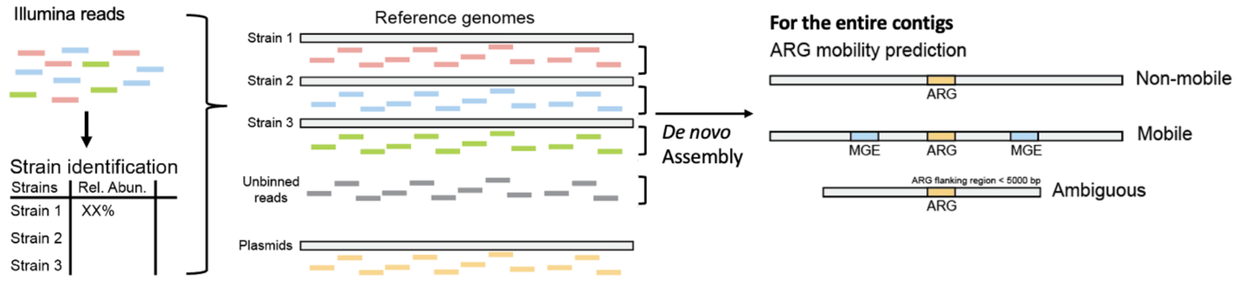


**Supplementary Fig. 11** Schematic of the integrated bioinformatics pipeline that combines *de novo* assembly with reference-based binning for ARG annotation and mobility prediction.

### Supplementary Tables

**Supplementary Table 1** Household drinking water survey data for enrolled households (*n* = 50) in Dagoretti South and Kibera

| Household drinking water | Overall (*n* = 50) | Dagoretti South (*n* = 25) | Kibera (*n* = 25) |
|---|---|---|---|
| Drinking water source types | | | |
| Borehole | 11 (22%) | 11 (44%) | 0 (0%) |
| Piped water into yard/plot | 5 (10%) | 0 (0%) | 5 (20%) |
| Piped water outside the compound | 31 (62%) | 11 (44%) | 20 (80%) |
| Tanker truck | 3 (6%) | 3 (12%) | 0 (0%) |
| Post water management | | | |
| Bottled chlorine | 3 (6%) | 1 (4%) | 2 (8%) |
| Boiling | 5 (10%) | 1 (4%) | 4 (16%) |
| Filter (ceramic, sand, composite) | 1 (2%) | 1 (4%) | 0 (0%) |

**Supplementary Table 2** Household assets survey data for enrolled households (*n* = 50) in Dagoretti South and Kibera

| Household assets | Overall (*n* = 50) | Dagoretti South (*n* = 25) | Kibera (*n* = 25) |
|---|---|---|---|
| Electricity | 50 (100%) | 25 (100%) | 25 (100%) |
| Radio | 40 (80%) | 23 (92%) | 17 (68%) |
| TV | 47 (94%) | 25 (100%) | 22 (88%) |
| Mobile | 49 (98%) | 25 (100%) | 24 (96%) |
| Clock | 14 (28%) | 5 (20%) | 9 (36%) |
| Bicycle | 9 (16%) | 7 (28%) | 2 (8%) |
| Motorcycle | 5 (10%) | 4 (16%) | 1 (4%) |
| Stove | 34 (68%) | 16 (64%) | 18 (72%) |
| Cooker | 40 (80%) | 22 (88%) | 18 (72%) |
| Car | 1 (2%) | 1 (4%) | 0 (0%) |

**Supplementary Table 3.** Household latrine survey data for enrolled households ($n$ = 50) in Dagoretti South and Kibera

| Household latrine | Overall ($n$ = 50) | Dagoretti South ($n$ = 25) | Kibera ($n$ = 25) |
|---|---|---|---|
| Location | | | |
| Inside compound | 24 (48%) | 19 (76%) | 5 (2%) |
| Immediately outside compound (< 5 m away) | 7 (14%) | 4 (16%) | 3 (12%) |
| Outside compound (> 5 m away) | 18 (36%) | 2 (8%) | 16 (64%) |
| No answer | 1 (2%) | 0 (0%) | 1 (4%) |

**Supplementary Table 4.** Household animal practice survey data for enrolled households (*n* = 50) in Dagoretti South and Kibera

| Household animal practice | Overall (*n* = 50) | Dagoretti South (*n* = 25) | Kibera (*n* = 25) |
|---|---|---|---|
| Purpose for poultry ownership | | | |
| Meat | 45 (90%) | 22 (88%) | 23 (92%) |
| Eggs | 39 (78%) | 22 (88%) | 17 (68%) |
| Income generation | 31 (62%) | 11 (44%) | 20 (80%) |
| Selling bird | 29 (58%) | 10 (40%) | 19 (76%) |
| Selling meat | 2 (4%) | 0 (0%) | 2 (8%) |
| Selling eggs | 7 (14%) | 6 (24%) | 1 (4%) |
| Pet | 1 (2%) | 0 (0%) | 1 (4%) |
| For gift to a visitor | 1 (2%) | 0 (0%) | 1 (4%) |
| Poultry and canine entering house | | | |
| Always / Often | 13 (26%) | 3 (12%) | 10 (40%) |
| Sometimes | 15 (30%) | 10 (40%) | 5 (20%) |
| Never | 22 (44%) | 12 (48%) | 10 (40%) |
| Feces near household soil sampling area | | | |
| Yes | 18 (36%) | 6 (24%) | 12 (48%) |

**Supplementary Table 5** Pairwise permutation test of mean strain-sharing rates among sample types within and between households.

| Comparison | Sharing type 1 | Sharing type 2 | Mean rate (Sharing type1) | Mean rate (Sharing type2) | *p* | adj. *p* |
|---|---|---|---|---|---|---|
| Within | Human-Animal | Animal-Household soil | 0.00166 | 0.02941 | 0.056 | 0.104 |
| | Human-Animal | Animal-Drinking water | 0.00166 | 0.00000 | 0.904 | 0.904 |
| | Human-Animal | Human-Household soil | 0.00166 | 0.00000 | 0.835 | 0.904 |
| | Human-Animal | Human-Drinking water | 0.00166 | 0.06463 | 0.003 | 0.027 |
| | Animal-Household soil | Animal-Drinking water | 0.02941 | 0.00000 | 0.042 | 0.104 |
| | Animal-Household soil | Human-Household soil | 0.02941 | 0.00000 | 0.180 | 0.270 |
| | Animal-Household soil | Human-Drinking water | 0.02941 | 0.06463 | 0.283 | 0.364 |
| | Animal-Drinking water | Human-Household soil | 0.00000 | 0.00000 | | - |
| | Animal-Drinking water | Human-Drinking water | 0.00000 | 0.06463 | 0.009 | 0.041 |
| | Human-Household soil | Human-Drinking water | 0.00000 | 0.06463 | 0.056 | 0.104 |
| Between | Human-Animal | Animal-Household soil | 0.00128 | 0.00214 | 0.403 | 0.734 |
| | Human-Animal | Animal-Drinking water | 0.00128 | 0.00089 | 0.625 | 0.781 |
| | Human-Animal | Human-Household soil | 0.00128 | 0.00123 | 0.944 | 0.944 |
| | Human-Animal | Human-Drinking water | 0.00128 | 0.00030 | 0.117 | 0.390 |
| | Animal-Household soil | Animal-Drinking water | 0.00214 | 0.00089 | 0.242 | 0.605 |
| | Animal-Household soil | Human-Household soil | 0.00214 | 0.00123 | 0.476 | 0.734 |
| | Animal-Household soil | Human-Drinking water | 0.00214 | 0.00030 | 0.105 | 0.390 |
| | Animal-Drinking water | Human-Household soil | 0.00089 | 0.00123 | 0.770 | 0.856 |
| | Animal-Drinking water | Human-Drinking water | 0.00089 | 0.00030 | 0.514 | 0.734 |
| | Human-Household soil | Human-Drinking water | 0.00123 | 0.00030 | 0.095 | 0.390 |

**Supplementary Table 6** Relative frequency of ARG clusters with predicted mobility (in a separate Excel sheet)

**Supplementary Table 7** Description of the simulated datasets for benchmarking

| | | GTEN 247 | GTEN 291 | GTEN 293 | GTEN 306 | GTEN 378 |
|---|---|---|---|---|---|---|
| | Genome size (Mbp) | 5.5 | 5.0 | 4.9 | 5.2 | 4.7 |
| Ratio1 | Pooled ratio | 1 | 1 | 1 | 1 | 1 |
| | Expected coverage | 73.6X | 80.6X | 81.3X | 77.5X | 84.4X |
| | StrainGST identification | *E. coli* TUM20902 | - | *E. coli* MB19 | *E. coli* 14EC033 | *E. coli* 2011C-3911 |
| | StrainGST relative abundance (%) | 15.5 | - | 24.6 | 28.3 | 24.9 |
| Ratio2 | Pooled ratio | 1 | 0.1 | 1 | 1 | 1 |
| | Expected coverage | 88.5X | 9.8X | 99.2X | 95.4X | 103.0X |
| | StrainGST identification | *E. coli* TUM20902 | - | *E. coli* MB19 | *E. coli* 14EC033 | *E. coli* 2011C-3911 |
| | StrainGST relative abundance (%) | 17.3 | - | 19.3 | 30.7 | 25.6 |
| Ratio3 | Pooled ratio | 1 | 0.025 | 1 | 1 | 1 |
| | Expected coverage | 90.2X | 2.5X | 101.0X | 96.3X | 104.9X |
| | StrainGST identification | *E. coli* TUM20902 | - | *E. coli* MB19 | *E. coli* 14EC033 | *E. coli* 2011C-3911 |
| | StrainGST relative abundance (%) | 17.5 | - | 18.9 | 30.9 | 25.6 |
| Ratio4 | Pooled ratio | 0.1 | 1 | 1 | 1 | 10 |
| | Expected coverage | 2.8X | 30.8X | 31.0X | 29.6X | 322.3X |
| | StrainGST identification | *E. coli* TUM20902 | - | *E. coli* MB19 | *E. coli* 14EC033 | *E. coli* 2011C-3911 |
| | StrainGST relative abundance (%) | 1.9 | - | 17.2 | 12.2 | 61.9 |
| Ratio5 | Pooled ratio | 0.1 | 0.1 | 1 | 1 | 10 |
| | Expected coverage | 3.0X | 3.3X | 33.3X | 31.8X | 346.1X |
| | StrainGST identification | *E. coli* TUM20902 | - | *E. coli* MB19 | *E. coli* 14EC033 | *E. coli* 2011C-3911 |
| | StrainGST relative abundance (%) | 1.7 | - | 13.1 | 12.2 | 66.1 |
| Ratio6 | Pooled ratio | 0.1 | 0.025 | 1 | 1 | 10 |
| | Expected coverage | 3.0X | 0.8X | 33.5X | 32.0X | 348.2X |
| | StrainGST identification | *E. coli* TUM20902 | - | *E. coli* MB19 | *E. coli* 14EC033 | *E. coli* 2011C-3911 |
| | StrainGST relative abundance (%) | 1.6 | - | 12.8 | 12.1 | 66.7 |

**Supplementary Table 8.** Characteristics of the isolates used for the simulated datasets

| Identifier | Genome size (Mbp) | Phylogroup | StrainGST identification | Number of plasmids | Number of unique ARG clusters | | |
|---|---|---|---|---|---|---|---|
| | | | | | Overall | Non-mobile | Mobile |
| GTEN 247 | 5.5 | D | *E. coli* TUM20902 | 4 | 57 | 30 | 27 |
| GTEN 291 | 5.0 | A | *E. coli* NCTC9087 | 1 | 46 | 25 | 21 |
| GTEN 293 | 4.9 | A | *E. coli* MB19 | 2 | 44 | 27 | 17 |
| GTEN 306 | 5.2 | E | *E. coli* 14EC033 | 3 | 47 | 19 | 28 |
| GTEN 378 | 4.7 | B1 | *E. coli* 2011C-3911 | 0 | 45 | 30 | 15 |
